# Glymphatic dysfunction across sleep disorders and its cognitive implications

**DOI:** 10.64898/2026.10.20.26366043

**Authors:** Ruth Whitehouse, Tracy D. Farr, Shen Li, Robert T. R. Huckstepp, Johannes Boltze

**Affiliations:** School of Life Sciences, University of Warwick, Coventry CV4 7AL, United Kingdom; MRC Cognition and Brain Sciences Unit, University of Cambridge, Cambridge CB2 7EF, United Kingdom; Institute for Neuroscience and Cardiovascular Research, Queen’s Medical Research Institute, University of Edinburgh, Edinburgh EH16 4TJ, United Kingdom; Department of Neurology and Psychiatry, Beijing Shijitan Hospital, Capital Medical University, Beijing, China; Beijing Institute of Brain Disorders, Capital Medical University, Beijing, China; The Montreal Neurological Institute-Hospital, McGill University, Montreal H3A 2B4, Canada

**Keywords:** Glymphatic system, sleep disturbance, obstructive sleep apnea, sleep deprivation, narcolepsy type 1, perivascular clearance, cognitive impairment, neuroimaging

## Abstract

Sleep disorders such as obstructive sleep apnea (OSA), sleep deprivation (SD), and narcolepsy type 1 (NT1), are associated with cognitive dysfunction and an increased risk of neurodegenerative disease. Glymphatic transport disruption has been suggested as one potential mechanistic link. However, evidence across sleep disorders and glymphatic-related metrics remains fragmented. We aimed to determine whether glymphatic-related metrics are altered in OSA, SD, and NT1. We performed a systematic review and meta-analysis of clinical and preclinical studies to determine whether glymphatic-related metrics are altered in OSA, SD, and NT1. Across studies of OSA, glymphatic-related metrics indicated significant impairment of glymphatic transport with a moderate negative association between apnea-hypopnea index (AHI) and diffusion tensor imaging along the perivascular space (DTI-ALPS) index (r=−0.53, 95% confidence interval (CI): −0.63 to −0.41; *p*<0.001). Available evidence did not demonstrate statistically significant alterations in glymphatic measures in SD or NT1. These findings suggest that impaired glymphatic transport may represent a potential pathway linking OSA to cognitive decline and neurodegenerative risk. However, current evidence is insufficient to determine whether comparable alterations occur in SD or NT1. Substantial heterogeneity across studies highlights the need for standardized reporting and harmonized methodologies to advance the field.

## 1. Introduction

The glymphatic system has been proposed as a brain-wide perivascular transport system facilitating cerebrospinal fluid (CSF)-interstitial fluid (ISF) exchange and solute clearance from the central nervous system (CNS)^1^. CSF enters the brain along periarterial spaces surrounding penetrating arteries, exchanges with ISF within the brain parenchyma, and exits along perivenous pathways^2^. Solutes transported through the glymphatic system are thought to ultimately drain via the meningeal lymphatic vessels to the deep cervical lymph nodes^3^. Water transport across astrocytic endfeet expressing aquaporin-4 (AQP4) channels has been proposed to facilitate perivascular exchange^4–6^. However, whether glymphatic transport meaningfully contributes to large-scale solute clearance in the human brain remains actively debated^7^. This ongoing debate partly reflects methodological limitations in current in vivo techniques used to quantify glymphatic transport, as well as their susceptibility to physiological and imaging-related confounds^8^.

Despite these uncertainties, increasing evidence suggests that perivascular transport pathways may contribute to the clearance of neurotoxic metabolites from the brain, including amyloid-β and tau, key proteins implicated in Alzheimer’s disease (AD) pathology^1,9^. Disruption of these clearance mechanisms may promote pathogenic protein accumulation within the brain parenchyma. In studies using animal models of AD, disruption of meningeal lymphatic drainage worsens amyloid pathology and accelerates cognitive decline in animal models of AD^10^. In humans, neuroimaging studies indicated alterations in glymphatic-related metrics in AD patients^11^. This suggests that impaired perivascular clearance may contribute to pathogenic protein accumulation and related downstream consequences.

Glymphatic transport is most efficient during sleep, particularly slow-wave sleep^12^. Glymphatic influx has also been shown to correlate with electroencephalogram (EEG) delta power, linking glymphatic transport to the intensity of slow-wave activity^13^. Mechanistically, reduced noradrenergic signaling during sleep is thought to promote expansion of the interstitial space, facilitating greater perivascular exchange and solute transport^12^. Additional evidence from human neuroimaging studies demonstrates that slow-wave activity is temporally coupled to large amplitude oscillations in CSF flow, suggesting that sleep architecture directly regulates CSF dynamics^14^. Collectively, these findings identify sleep as a key physiological regulator of glymphatic transport. Disruption of sleep architecture may therefore impair glymphatic transport.

Several common sleep disorders, including narcolepsy type 1 (NT1), sleep deprivation (SD), and obstructive sleep apnea (OSA), are characterized by fragmentation of sleep architecture and reductions in slow-wave sleep^15,16^ and are associated with cognitive impairment. Epidemiological studies further linked OSA and chronic SD with cognitive decline and dementia^17–19^. However, the biological mechanisms remain poorly understood, and it is unclear whether glymphatic dysfunction may mediate or contribute to these associations.

Investigating glymphatic function in humans is challenging as glymphatic function cannot be measured directly. Instead, most studies rely on indirect imaging proxies capturing different aspects of perivascular or CSF dynamics. Consequently, the evidence base is highly fragmented and methodologically heterogeneous, complicating the interpretation and synthesis of existing findings.

This systematic review and meta-analysis synthesizes preclinical and human studies examining alterations in glymphatic-related metrics across OSA, SD, and NT1. By integrating findings across multiple classes of glymphatic-related measurements, this study aims to investigate whether sleep disturbances are associated with impaired glymphatic transport. A secondary objective is to examine whether these sleep disturbances are also associated with cognitive dysfunction, providing insight into potential mechanistic links between sleep disturbance, impaired clearance pathways, and cognitive decline.

## 2. Methods

### 2.1 Study selection

A systematic review and meta-analysis were conducted in accordance with the Preferred Reporting Items for Systematic Reviews and Meta-analyses (PRISMA) 2020 guidelines^20^. The literature search included preclinical and clinical studies. We also assessed study quality, risk of bias, and robustness of assessment. A detailed description of the search approach, data extraction and statistical analysis are available in the supplementary methods and Supplementary Tables 1 to 5.

## 3. Results

### 3.1 Study selection

Searches yielded 1,168 records, which were reduced to 873 after removal of duplicates. Following title and abstract screening, 812 records were excluded, and 61 full text articles were assessed for eligibility. Of these, 41 were excluded because they did not report a direct measure of glymphatic function, but instead reported other biomarkers (e.g., amyloid-β and tau) as proxies for clearance. The study selection process is summarized in Figure 1.

**Figure 1.**
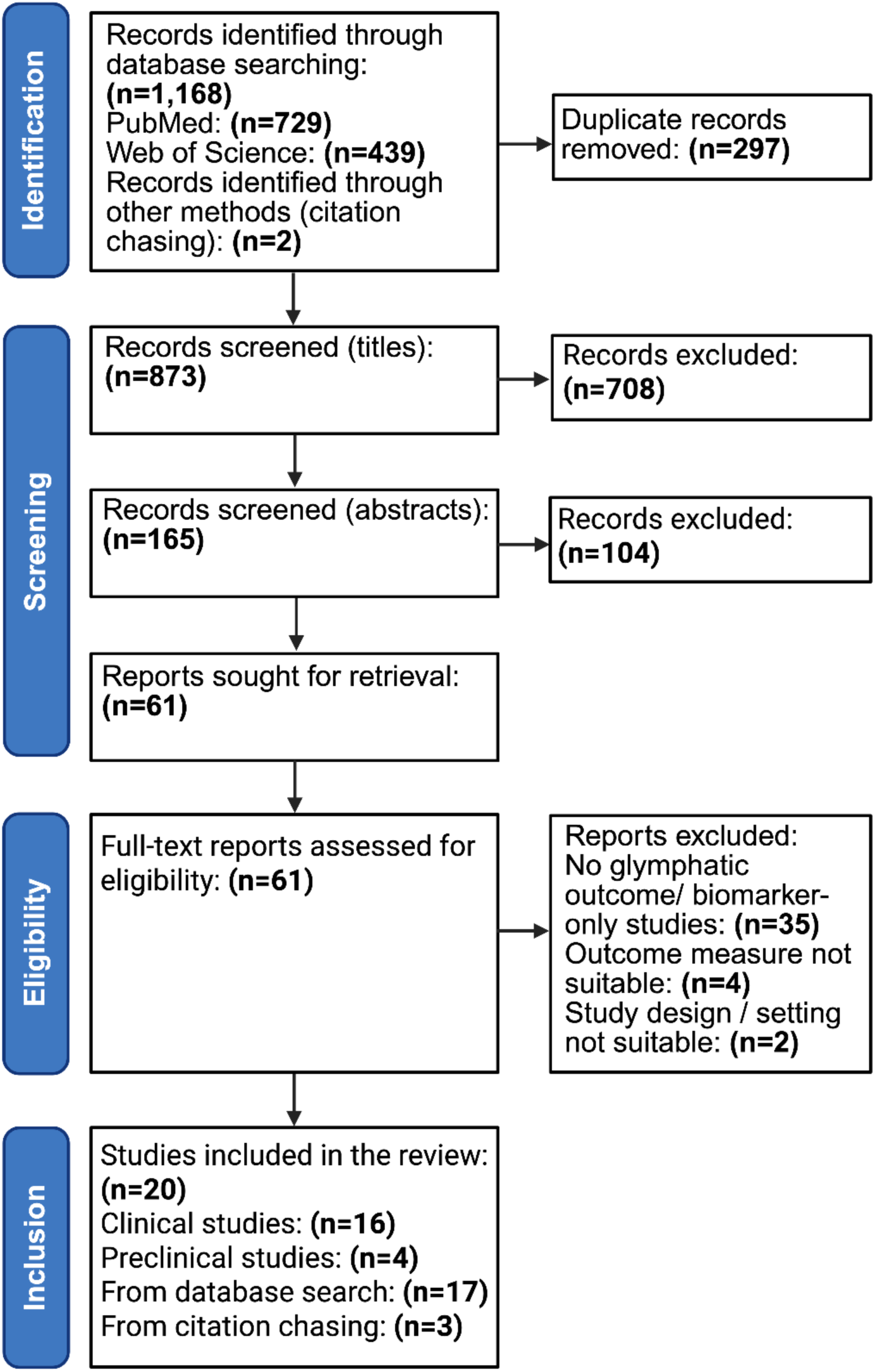
PRISMA flow diagram of the study selection process.

The final pool of eligible studies included 20 studies (16 clinical; four preclinical). Across the 16 clinical studies, a total of 1,095 participants were included. Of the 20 included studies, 12 clinical studies contributed data to the meta-analysis. The remaining eight studies (four clinical and four preclinical studies) were synthesized narratively. This was due to insufficient quantitative data in one clinical OSA study and substantial methodological heterogeneity across the remaining three clinical NT1 and the four preclinical studies. The main characteristics of studies included in the meta-analysis are summarized in Table 1, with further demographic and methodological details provided in Supplementary Table 6. Characteristics of studies included in the narrative synthesis are presented in Table 2, with additional study information detailed in Supplementary Table 6.

**Table 1.** Main study characteristics of included meta-analysis studies. Abbreviations: HC, healthy controls; OSA, obstructive sleep apnea; SD, sleep deprivation; DTI-ALPS, diffusion tensor image analysis along the perivascular space; CP, choroid plexus; ICV, intracranial volume; D*, pseudodiffusion coefficient; MoCA, Montreal Cognitive Assessment; GPCOG, General Practitioner Assessment of Cognition; MIS, Memory Impairment Screen.

| Author,<br>year | Study design | Population/<br>model | Sample<br>sizes | | Age (mean ±<br>Standard<br>Deviation)<br>(years) | | Sex (%<br>males) | | Sleep<br>disturbance<br>type | Glymphatic<br>outcome | Cognitive<br>outcome(s) | Main outcome(s) in<br>sleep disturbance vs<br>controls |
| --- | --- | --- | --- | --- | --- | --- | --- | --- | --- | --- | --- | --- |
|  |  |  | HC | SD | HC | SD | HC | SD |  |  |  |  |
| Lin et al.,<br>2024<sup>22</sup> | Cross-sectional<br>case-control | Adults with<br>OSA vs. HC | 50 | 84<sup>†</sup> | 37.0 ±<br>8.6 | 33.4<br>±<br>8.9<sup>†</sup> | 66.0 | 76.2<sup>†</sup> | OSA | DTI-ALPS<br>index (bilateral<br>mean) | Stroop test | Significantly reduced<br>DTI-ALPS index;<br>significantly increased<br>Stroop completion time. |
| Lee et al.,<br>2022<sup>23</sup> | Cross-sectional<br>case-control | Adults with<br>OSA vs. HC | 24 | 24 | 63.7 ±<br>6.3 | 62.0<br>±<br>15.0 | 58.3 | 62.5 | OSA | DTI-ALPS<br>index (bilateral<br>mean) | None | Significantly reduced<br>DTI-ALPS index. |
| Xiong et al.,<br>2024<sup>21</sup> | Cross-sectional<br>case-control | Adults with<br>OSA vs. HC | 34 | 31 | 43.21<br>± 8.5 | 39.9<br>± 9.3 | 85.3 | 90.3 | OSA | DTI-ALPS<br>index (bilateral<br>mean) | MoCA | Significantly reduced<br>DTI-ALPS index;<br>significantly reduced<br>MoCA. |
| Zhao et al.,<br>2025<sup>61</sup> | Randomized<br>controlled trial<br>(parallel-group) | Healthy adults<br>randomized to<br>SD vs. control sleep | 13 | 51 | 36.4 ±<br>10.3 | 33.1<br>± 8.7 | 46.2 | 58.8 | Total SD (24<br>h) | gBOLD-CSF<br>coupling<br>strength | None | Significantly<br>strengthened gBOLD-CSF coupling. |
| Custer et al.,<br>2025<sup>76</sup> | Randomized<br>controlled trial<br>(parallel-group) | Healthy adults<br>randomized to<br>SD vs. control<br>sleep | 60 | 56 | NR | NR | 52.0 | 50.0 | Partial SD<br>(24 h; <4 h<br>sleep) | Centrum<br>Semiovale<br>PVS volume<br>fraction<br>(reported in<br>mm<sup>3</sup>) | None | No significant difference<br>in PVS volume fraction. |
| Kim et al.,<br>2024<sup>77</sup> | Cross-sectional<br>case-control | Adults with<br>OSA vs. HC | 38 | 41 | 61.7 ±<br>6.8 | 62.7<br>±<br>13.9 | 73.6 | 62.5 | OSA | CP volume<br>(ICV-corrected, %) | None | Significantly increased<br>CP volume. |
| Eide and<br>Ringstad,<br>2021<sup>78</sup> | Non-randomized<br>controlled<br>intervention<br>(parallel-group) | Adults<br>allocated to<br>SD vs. control<br>sleep | 9 | 7 | 36.1 ±<br>13.0 | 44.7<br>±<br>15.7 | 11.1 | 14.3 | Total SD (24<br>h) | Tracer<br>enrichment in<br>parasagittal<br>dura at 24 h | None | No significant difference<br>in parasagittal dura<br>tracer enrichment. |
| Eide et al.,<br>2021<sup>79</sup> | Non-randomized<br>controlled<br>intervention<br>(parallel-group) | Adults<br>allocated to<br>SD vs. control<br>sleep | 17 | 7 | 39.2 ±<br>14.1 | 44.7<br>±<br>15.7 | 11.8 | 14.3 | Total SD (24<br>h) | Cerebral white<br>matter tracer<br>retention at 48<br>h (% change<br>T1 signal) | None | Significantly increased<br>white matter tracer<br>retention. |
| Li et al.,<br>2025<sup>28</sup> | Cross-sectional<br>case-control | Adults with<br>OSA vs. HC | 64 | 89 | 27.5 ±<br>12.5 | 31.8<br>± 8.0 | 78.1 | 81.1 | OSA | gBOLD-CSF<br>coupling<br>strength | None | Significantly weakened<br>gBOLD-CSF coupling. |
| Rane<br>Levendovsky<br>et al., 2024<sup>80</sup> | Randomized<br>controlled<br>crossover trial | Adults<br>completed<br>both normal<br>sleep and SD<br>(randomised<br>order) | 9 | 9 | 23.0<br>± 3.0 | 23.0<br>± 3.0 | 33.3 | 33.3 | Total SD (24<br>h) | D* | GPCOG and<br>MIS | Significantly reduced<br>D*; no significant<br>difference in GPCOG or<br>MIS. |
| Vinje et al.,<br>2023<sup>81</sup> | Non-randomized<br>controlled<br>intervention<br>(parallel-group) | Adults<br>allocated to<br>SD vs. control<br>sleep | 17 | 7 | 39.2 ±<br>14.1 | 44.7<br>±<br>15.7 | 11.8 | 12.5 | Total SD (24<br>h) | Brain-wide<br>tracer<br>retention at 48<br>h | None | Significantly increased<br>brain-wide tracer<br>retention. |
| Roy et al.,<br>2022<sup>82</sup> | Cross-sectional<br>case-control | Adults with<br>OSA vs. HC | 62 | 59 | 50.1 ±<br>10.4 | 49.9<br>±<br>10.0 | 54.8 | 59.3 | OSA | DTI-ALPS<br>index (bilateral<br>mean) | None | Significantly reduced<br>DTI-ALPS index. |

†Demographic characteristics (age and sex) for the sleep disturbance group were reported for the full experimental cohort, which included participants classified as snorers. Age and sex were not reported separately for the diagnosed OSA subgroups; therefore, the pooled sleep disturbance demographics (including snorers) are presented. The sleep disturbance sample size shown reflects only diagnosed OSA participants included in the meta-analysis (snorers excluded).

**Table 2.**
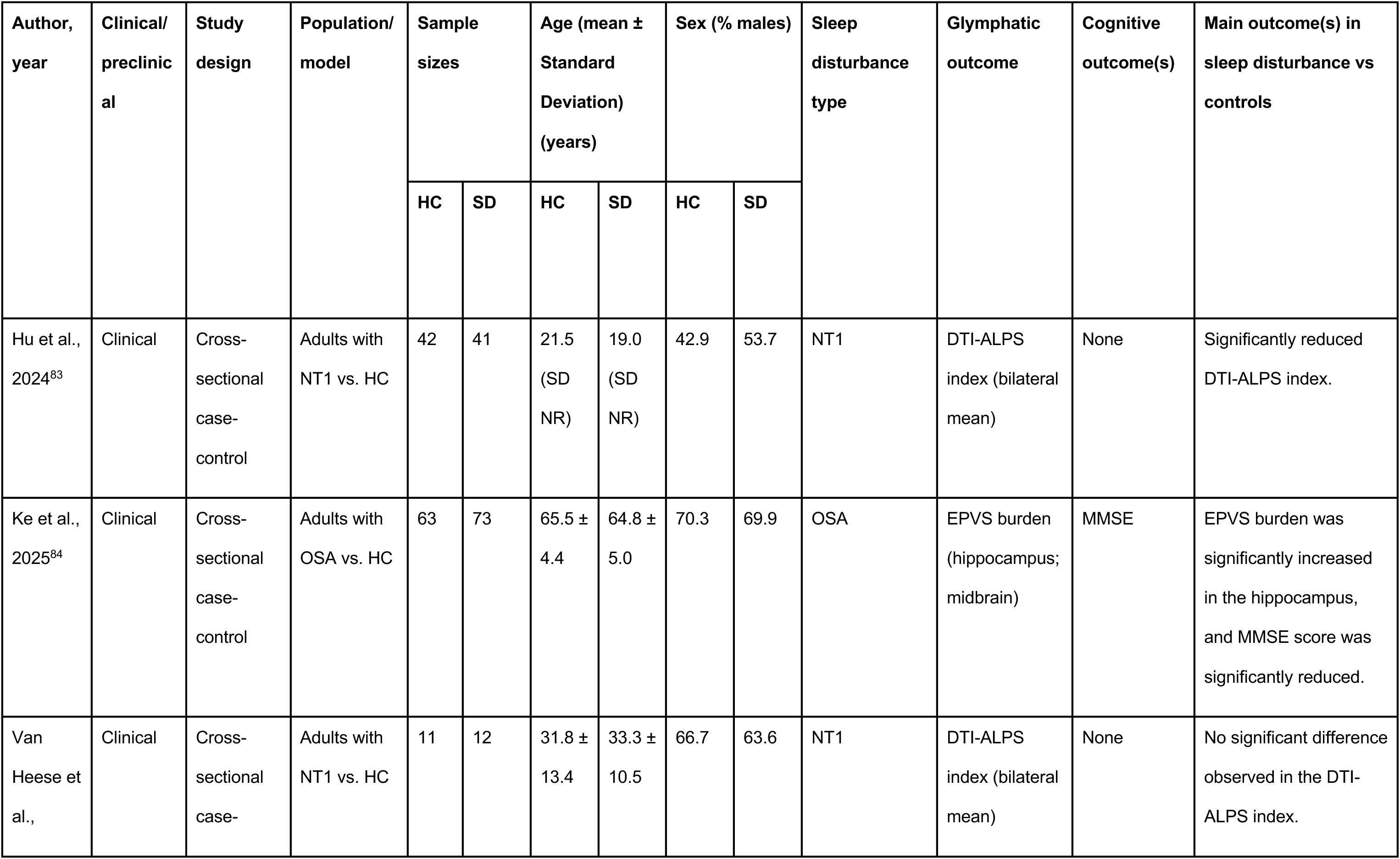
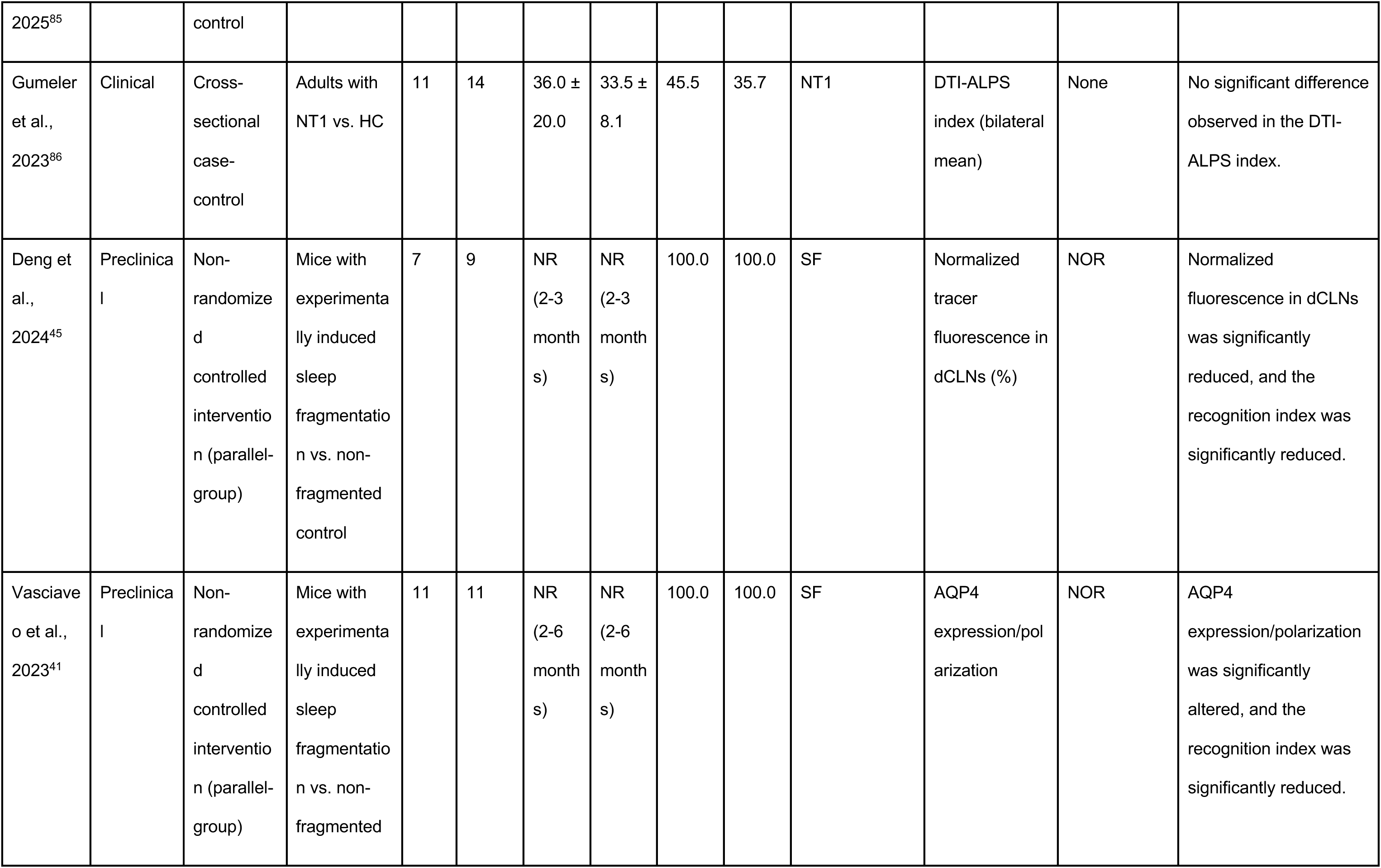
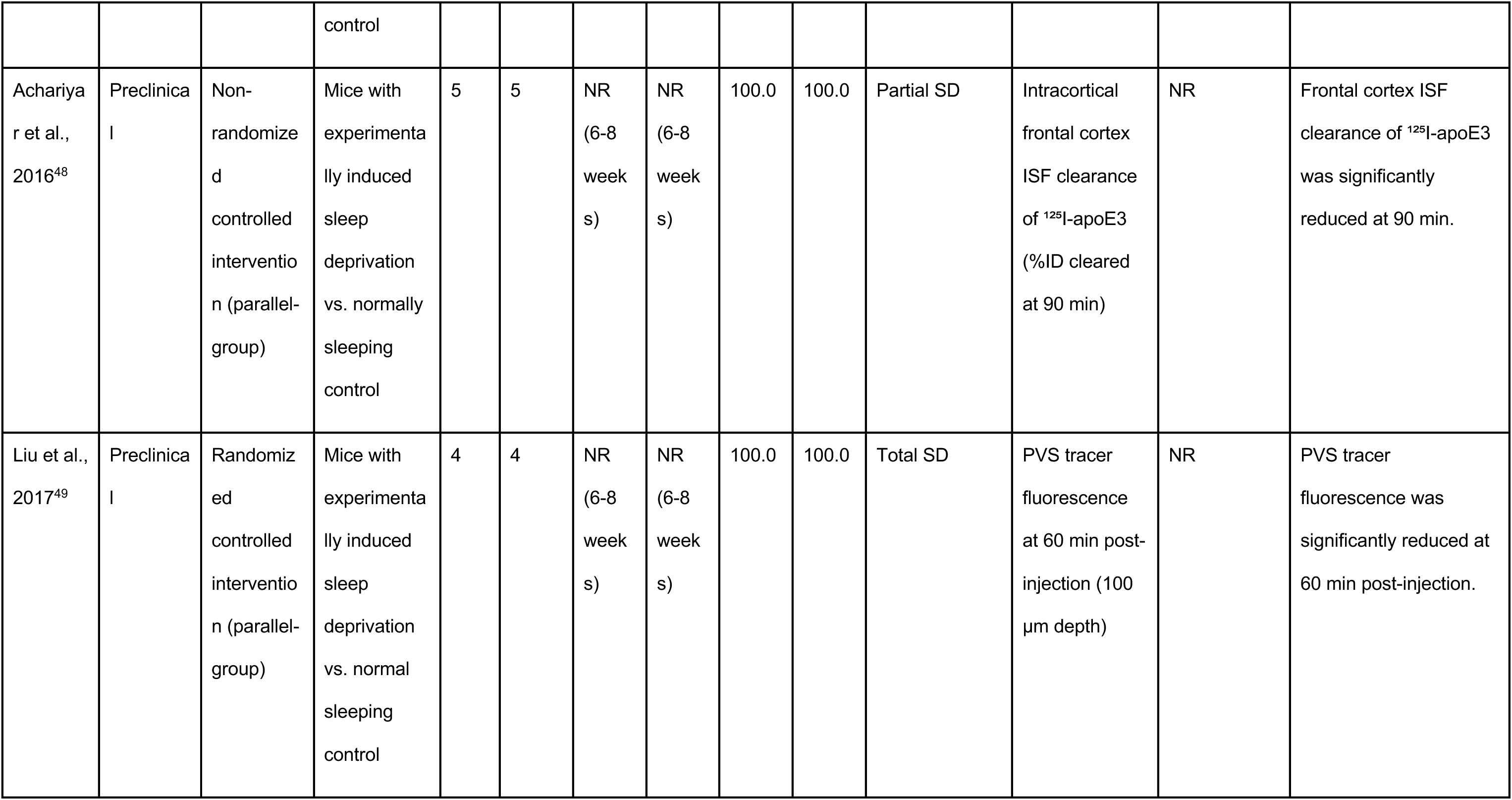
Main study characteristics of narratively synthesized studies. Abbreviations: HC, healthy controls; NT1, narcolepsy type 1; DTI-ALPS, diffusion tensor imaging analysis along the perivascular space; OSA, obstructive sleep apnea; EPVS, enlarged perivascular spaces; MMSE, Mini-Mental State Examination; SF, sleep fragmentation; dCLNs, deep cervical lymph nodes; NOR, Novel Object Recognition; AQP4, aquaporin-4; SD, sleep deprivation; %ID, percent injected dose; ISF, interstitial fluid; ¹²⁵I, iodine-125; NR, not reported; PVS, perivascular spaces; CSF, cerebrospinal fluid.

The clinical studies comprised ten case-control studies and six interventional studies with experimentally-induced sleep disruption, enrolling adults with varying severities and durations of sleep disorders. Studies were conducted in China (n=6), Norway (n=3), Korea, the USA (n=2 each), the Netherlands, Turkey, and Sweden (n=1 each). All included studies were published between 2021 and 2025, most frequently in 2024 and 2025 (n=5 each).

The four preclinical studies investigated male rodents, predominantly C57BL/6 mice (n=3), using models of sleep fragmentation or sleep deprivation (n=2 each). The age at first exposure to sleep disturbance ranged from 6 weeks to 6 months, and the duration of sleep disruption ranged from 3 hours to 30 days. Three studies used tracer-based metrics to assess glymphatic function, and one study explored AQP4 polarization as a proxy marker.

### 3.2 Clinical evidence: glymphatic consequences of sleep disorders

#### 3.2.1 Sleep deprivation

Six studies (137 sleep-deprived participants and 125 HC) evaluated the effects of sleep deprivation on glymphatic function (Figure 2A). The meta-analysis identified no statistically significant association between sleep deprivation and glymphatic measures (standardized mean difference (SMD)= −0.33; 95% CI: −0.79 to 0.13; p=0.12).

**Figure 2.**
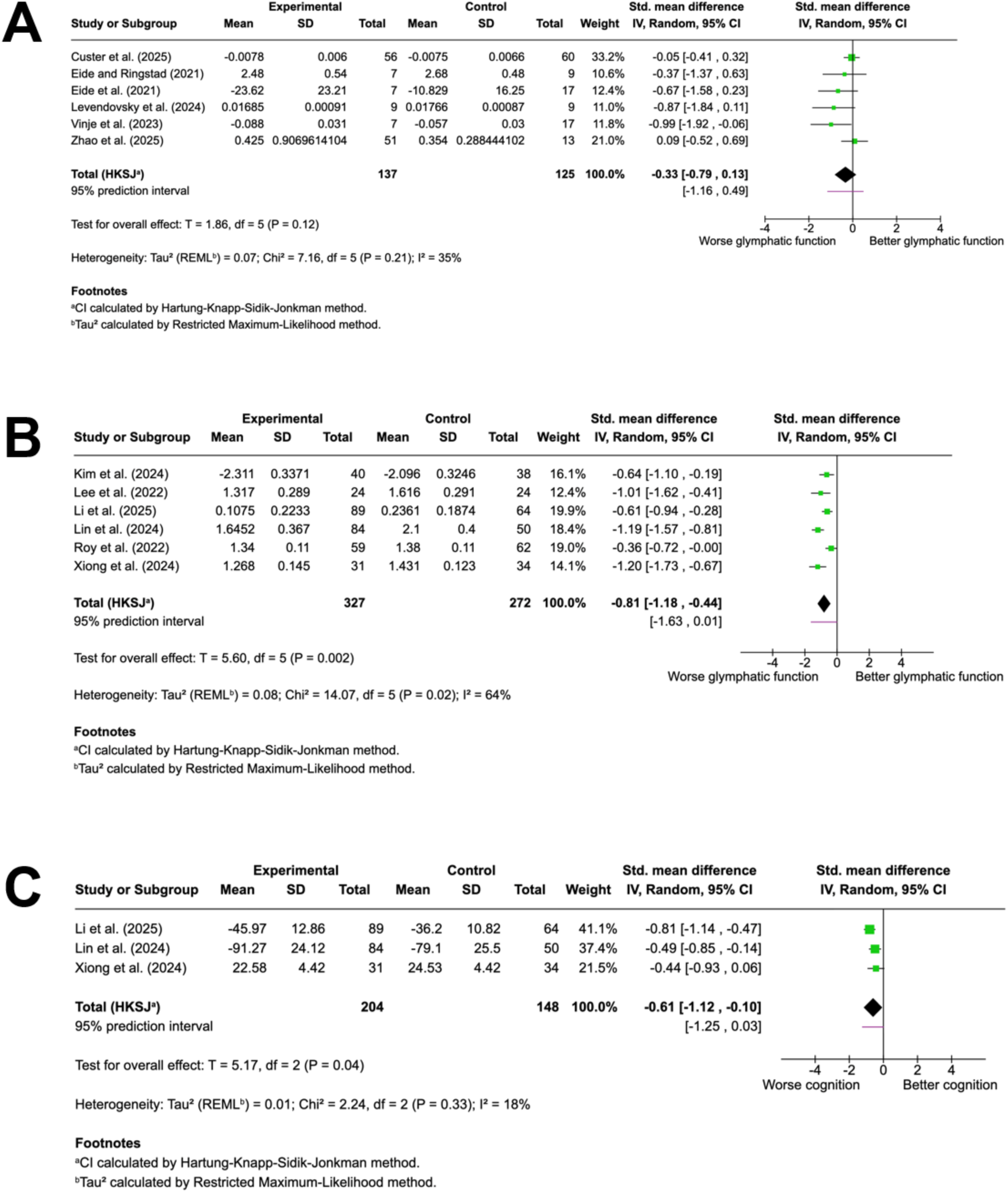
Forest plots summarizing clinical evidence for the effect of sleep deprivation and obstructive sleep apnea (OSA) on glymphatic function and cognitive performance. A) Sleep deprivation and glymphatic function. B) OSA and glymphatic function. C) OSA and cognition. Pooled standardized mean differences are shown with 95% confidence intervals and 95% prediction intervals.

Three sleep deprivation studies reported impaired glymphatic-related measures, two showed no effect, and one study reported an increase. Tracer MRI was mixed (two impaired clearance, one neutral). The increased measure came from an fMRI study reporting stronger BOLD-CSF coupling strength following sleep loss. IVIM suggested reduced microvascular-CSF exchange, while structural MRI showed no change in PVS volume fraction, indicating modality-dependent heterogeneity.

#### 3.2.2 Narcolepsy type 1

Three studies evaluated glymphatic-related metrics in NT1 but were summarized narratively due to limited and heterogeneous data. One study reported a significant reduction in DTI-ALPS index and enlarged choroid plexus volume, indicating impaired glymphatic function. However, two studies reported no significant differences in DTI-ALPS index or BOLD-CSF coupling strength compared with HC. Overall, the data is inconclusive and does not show a consistent impairment of glymphatic function in NT1.

#### 3.2.3 Obstructive sleep apnea

Six studies investigating the impact of OSA on glymphatic function (327 OSA patients and 272 HC) were included. The meta-analysis (Figure 2B) demonstrated a large and statistically significant impairment of glymphatic clearance in OSA compared with HC (SMD=−0.81, 95% CI: −1.18 to −0.44; p=0.002).

Across the seven narratively summarized clinical OSA studies, a consistent impairment of glymphatic function across several MRI-derived metrics was demonstrated. Four studies reported reduced DTI-ALPS index, one reported higher EPVS burden, one reported enlarged choroid plexus volume, and one reported reduced BOLD-CSF coupling strength. Five studies reported associations between the OSA severity (AHI) and glymphatic-related measures.

### 3.3 Cognitive consequences of obstructive sleep apnea

Three of the clinical OSA studies (204 OSA patients and 148 HC) reported cognitive outcomes, assessed using three neuropsychological tests: the Montreal Cognitive Assessment (MoCA), the Stroop test (total completion time), and a line tracing test. The pooled analysis demonstrated a significant reduction in cognitive performance associated with OSA compared with HC (SMD=−0.61; 95% CI: −1.12 to −0.10; p=0.04) (Figure 2C). However, this finding was sensitive to the inclusion of individual studies (see Section 3.5 for details).

### 3.4 Severity-dependent impairment in obstructive sleep apnea

Four studies discriminated OSA severity, with mild, moderate, and severe OSA groups compared with HC. However, Xiong et al^21^. did not report data for a mild OSA subgroup; therefore, the mild OSA analysis included three studies, whereas the severe and moderate analyses each included four studies. Mild OSA showed a non-significant effect (SMD=−0.61; 95% CI: −2.25 to 1.04; p=0.25). Heterogeneity in the mild OSA subgroup analysis was very high (I²=85%), indicating a substantial variability in effect sizes across studies (Figure 3A). The pooled effect for moderate OSA bordered on statistical significance (SMD=−0.83; 95% CI: −1.81 to 0.15; p=0.07), and high between-study heterogeneity (I²=73%) (Figure 3B). Severe OSA showed a large and statistically significant impairment in glymphatic clearance compared with HC (SMD=−1.23; 95% CI: −1.97 to −0.49; p=0.01), and moderate heterogeneity across studies (I²=61%) (Figure 3C). These findings may support a dose-response relationship between OSA severity and glymphatic dysfunction, with the largest effects seen in severe OSA.

**Figure 3.**
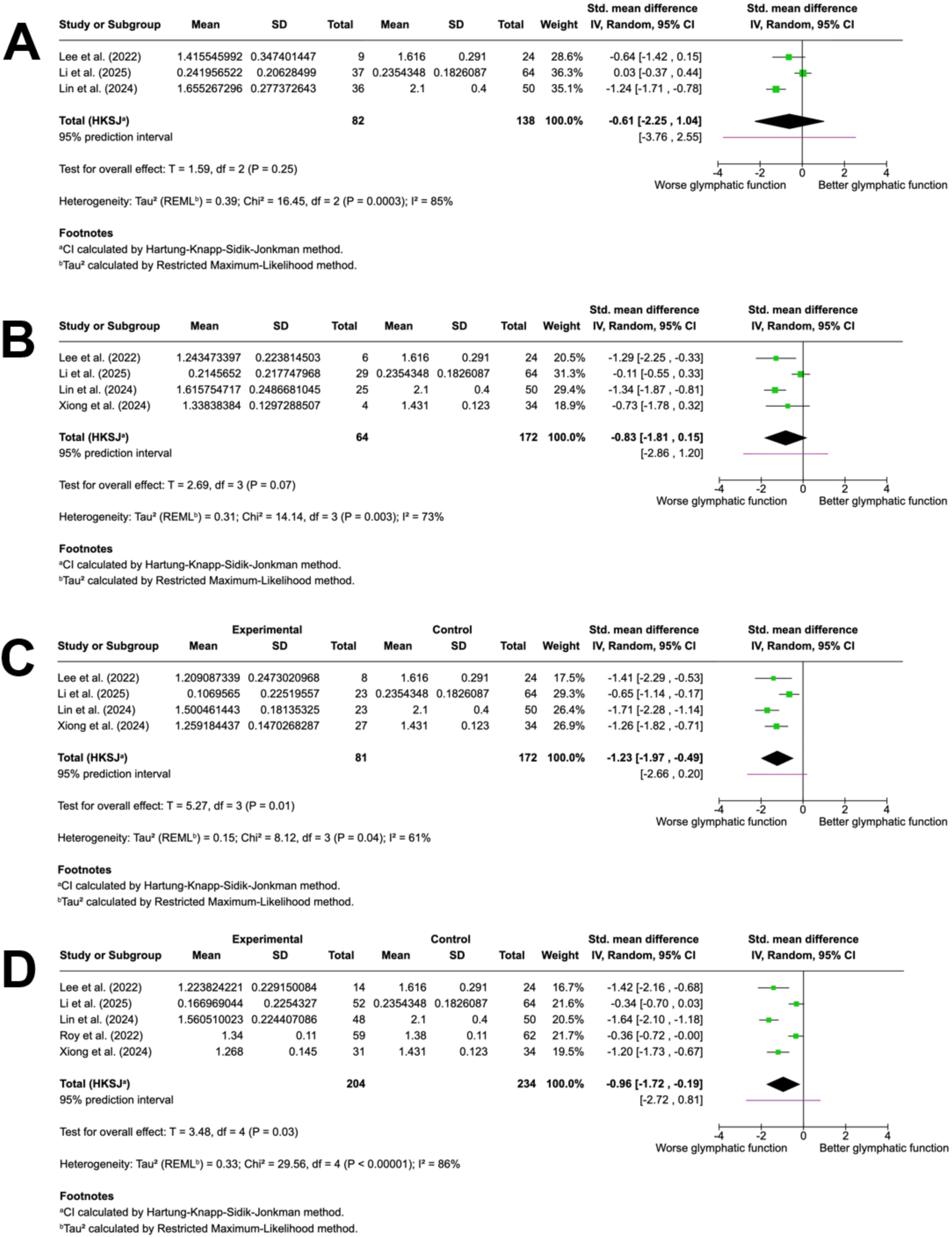
Forest plots summarizing clinical evidence for differences in glymphatic function between healthy controls and obstructive sleep apnea (OSA) groups stratified by severity. Panels show comparisons for (A) mild, (B) moderate, (C) severe OSA, and (D) combined moderate-severe OSA groups. OSA severity was classified according to the apnea-hypopnea index (AHI) thresholds. Pooled standardized mean differences are shown with 95% confidence intervals and 95% prediction intervals. The mild OSA analysis included three studies because Xiong et al. did not report data for a mild OSA subgroup. The moderate-severe OSA group analysis included five studies reporting either combined moderate-severe OSA data or sufficient subgroup data to allow aggregation of moderate and severe OSA subgroups.

As an additional exploratory analysis, studies reporting data that allowed moderate and severe OSA groups to be combined were pooled to assess whether restricting the analysis to higher-severity OSA altered the estimated effect. The analysis included five studies and showed reduced glymphatic-related measures in participants with moderate-severe OSA compared with HC (SMD=−0.96; 95% CI: −1.27 to −0.19; p=0.03) (Figure 3D). This effect estimate was slightly larger than that observed in the primary OSA analysis.

To further examine the relationship between OSA severity and glymphatic function as continuous variables, three studies were included in a correlation analysis. Individual data points for the diffusion tensor imaging along the PVS index (DTI-ALPS index) and the apnea-hypopnea index (AHI) were extracted, resulting in a combined sample of 159 participants (Lin et al., n=105^22^; Lee et al., n=23^23^; Xiong et al., n=31^21^).

The pooled correlation showed a moderate to strong inverse association between the DTI-ALPS index and AHI, consistent with a severity-dependent impairment in glymphatic function (r=−0.53; 95% CI: −0.63 to −0.41; *p*<0.001) (Figure 4A).

**Figure 4.**
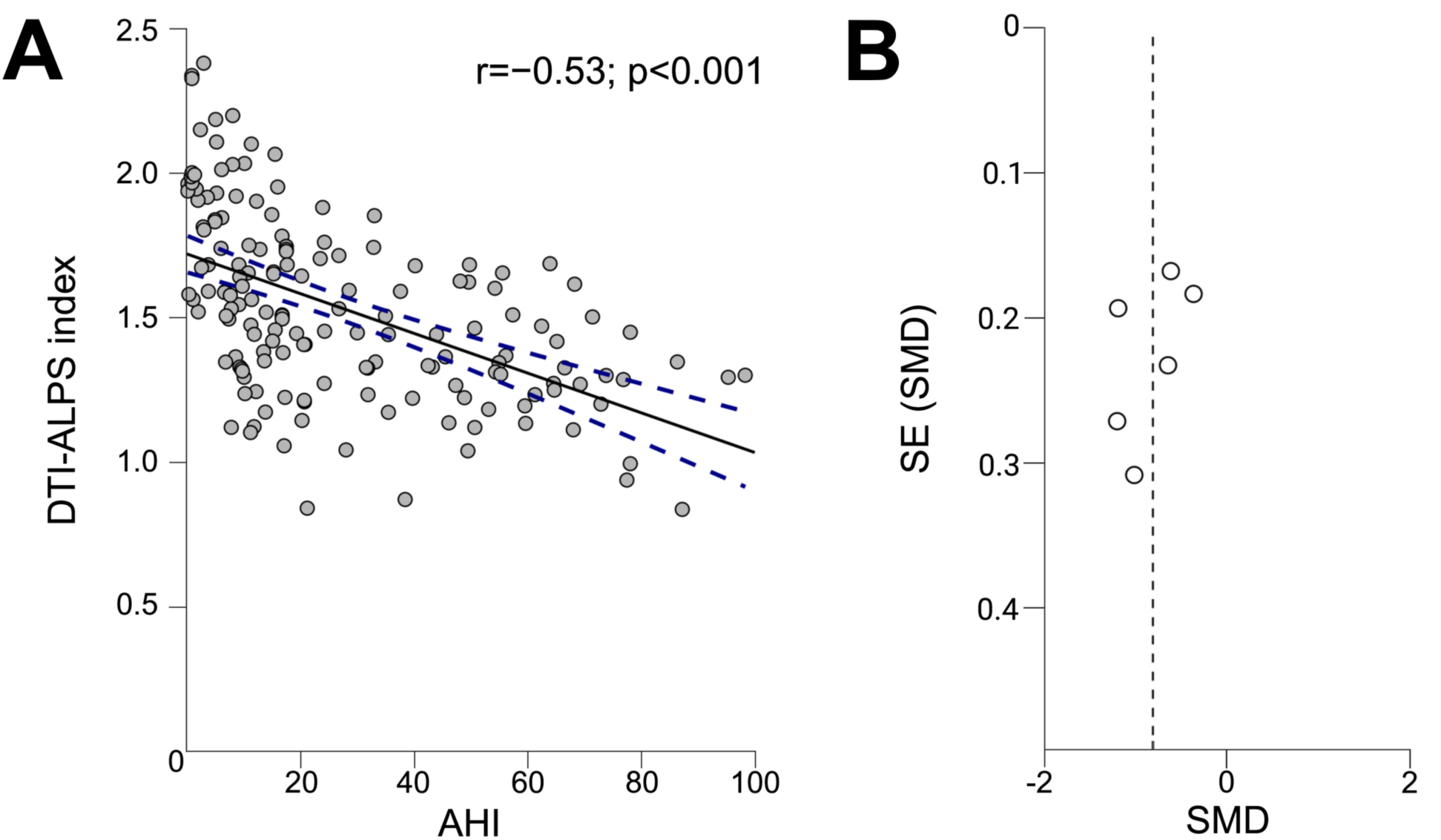
Relationship between obstructive sleep apnea (OSA) severity and glymphatic function, and assessment of publication bias. A) Correlation between Diffusion Tensor Imaging Along the Perivascular Space (DTI-ALPS) index and Apnea-Hypopnea Index (AHI), demonstrating an inverse association consistent with a severity-dependent impairment in glymphatic function. B) Funnel plot of clinical OSA studies with glymphatic outcomes compared with healthy controls, showing approximate symmetry, consistent with no evidence of publication bias.

### 3.5 Robustness of evidence

To evaluate the robustness of the principal meta-analytic findings, leave-one-out sensitivity analyses were conducted for OSA-related glymphatic outcomes and cognitive outcomes. For OSA-related glymphatic outcomes, the leave-one-out sensitivity analysis supported the robustness of the association; no single study disproportionately influenced the overall effect size (p-values 0.002 to 0.009). However, the analysis of OSA-related cognitive outcomes indicated greater fragility, with the statistical significance of the pooled effect size dependent on the inclusion of individual studies (p-values 0.04 to 0.17) (Table 3).

**Table 3.** Leave-one-out sensitivity analysis of cognitive performance outcomes in obstructive sleep apnea compared with healthy controls, demonstrating fragility of the pooled effect size.

| Omitted study | Hedges' *g* | 95% CI | p value |
| --- | --- | --- | --- |
| All studies included | -0.61 | -1.12 to -0.10 | 0.04 |
| Li et al., 2025<sup>28</sup> | -0.47 | -0.80 to -0.14 | 0.04 |
| Lin et al., 2024<sup>22</sup> | -0.67 | -2.95 to 1.61 | 0.17 |
| Xiong et al., 2024<sup>21</sup> | -0.65 | -2.66 to 1.35 | 0.15 |

Potential publication bias and small-study effects were assessed using funnel plots and Egger’s regression test. A funnel plot was generated only for OSA-related glymphatic outcomes, as this was the only analysis with a sufficient number of included studies to allow meaningful assessment of publication bias. The funnel plot showed approximate symmetry on visual inspection (Figure 4B). This was supported by non-significant Egger’s regression (t=−0.56; p=0.606), showing no evidence of small-study effects or publication bias, although the small number of included studies limits the power of this assessment.

### 3.6 Preclinical studies

Four preclinical studies were included: two using sleep fragmentation models and two using sleep deprivation models. Across studies, glymphatic-related transport was most commonly assessed using tracer-based methods (n=3), and AQP4-related outcomes were reported in two studies.

Across the three tracer-based studies, all reported reductions in glymphatic-related tracer influx and/or solute clearance following sleep disruption. In the tracer-based sleep fragmentation study, chronic fragmentation was associated with reduced tracer signals in the brain and downstream drainage compartments, whereas an acute fragmentation paradigm showed no significant effect. In sleep deprivation models, tracer-based outcomes reported reduced CSF/PVS tracer influx and/or reduced ISF solute clearance following sleep deprivation. One tracer study additionally reported isoform-dependent effects on apoE transport, with greater impairment for apoE4 than for apoE2 and apoE3.

AQP4-related outcomes indicated altered perivascular AQP4 organization following sleep disruption. Two sleep fragmentation studies assessed cognitive function, reporting a significant reduction in novel-object recognition performance in sleep fragmented mice.

Overall study quality was moderate (SYRCLE scores 8-10/16). Common methodological limitations included small sample sizes, incomplete reporting of experimental endpoints, and the absence of a priori power or sample size calculations (Table 4).

**Table 4.**
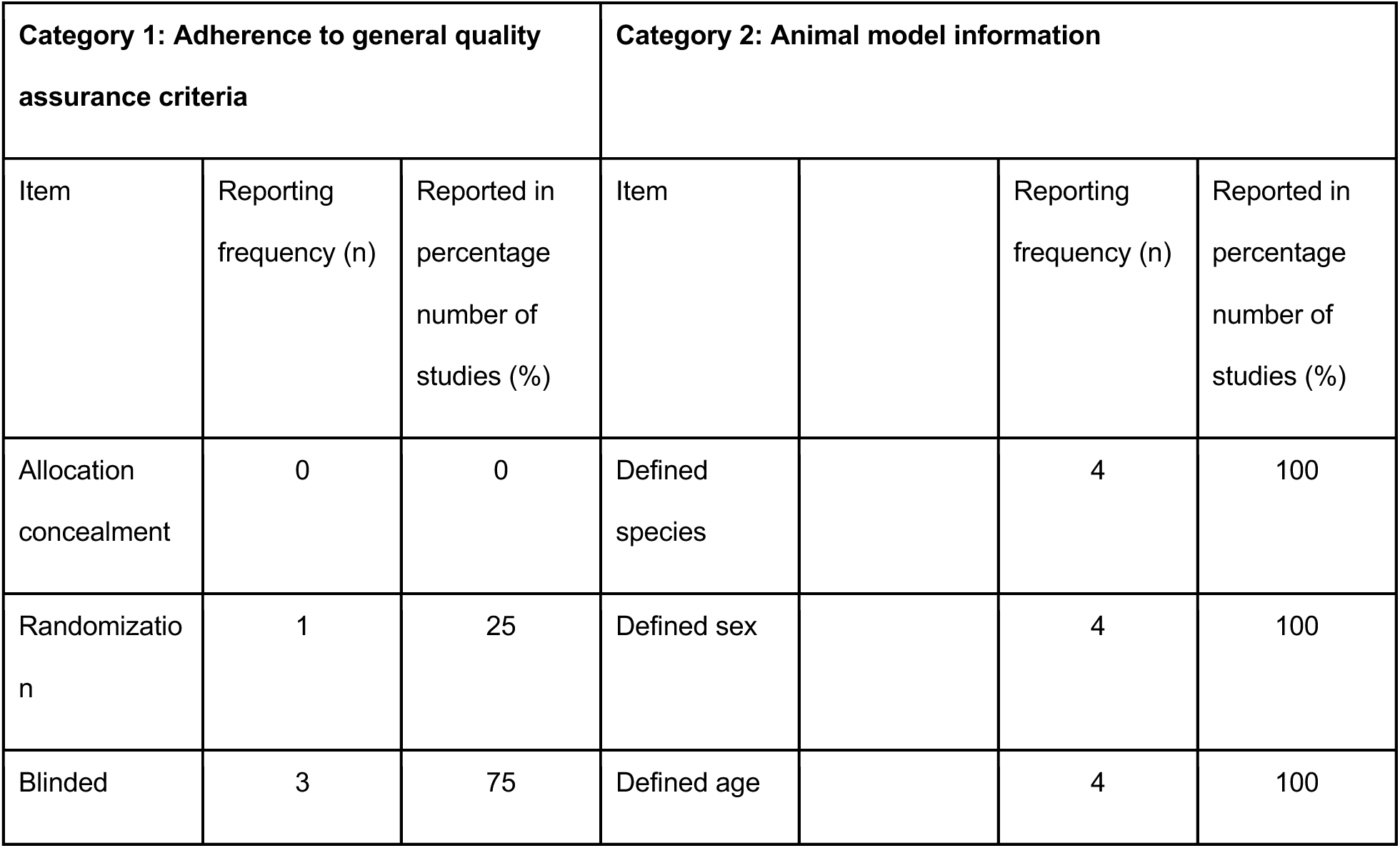
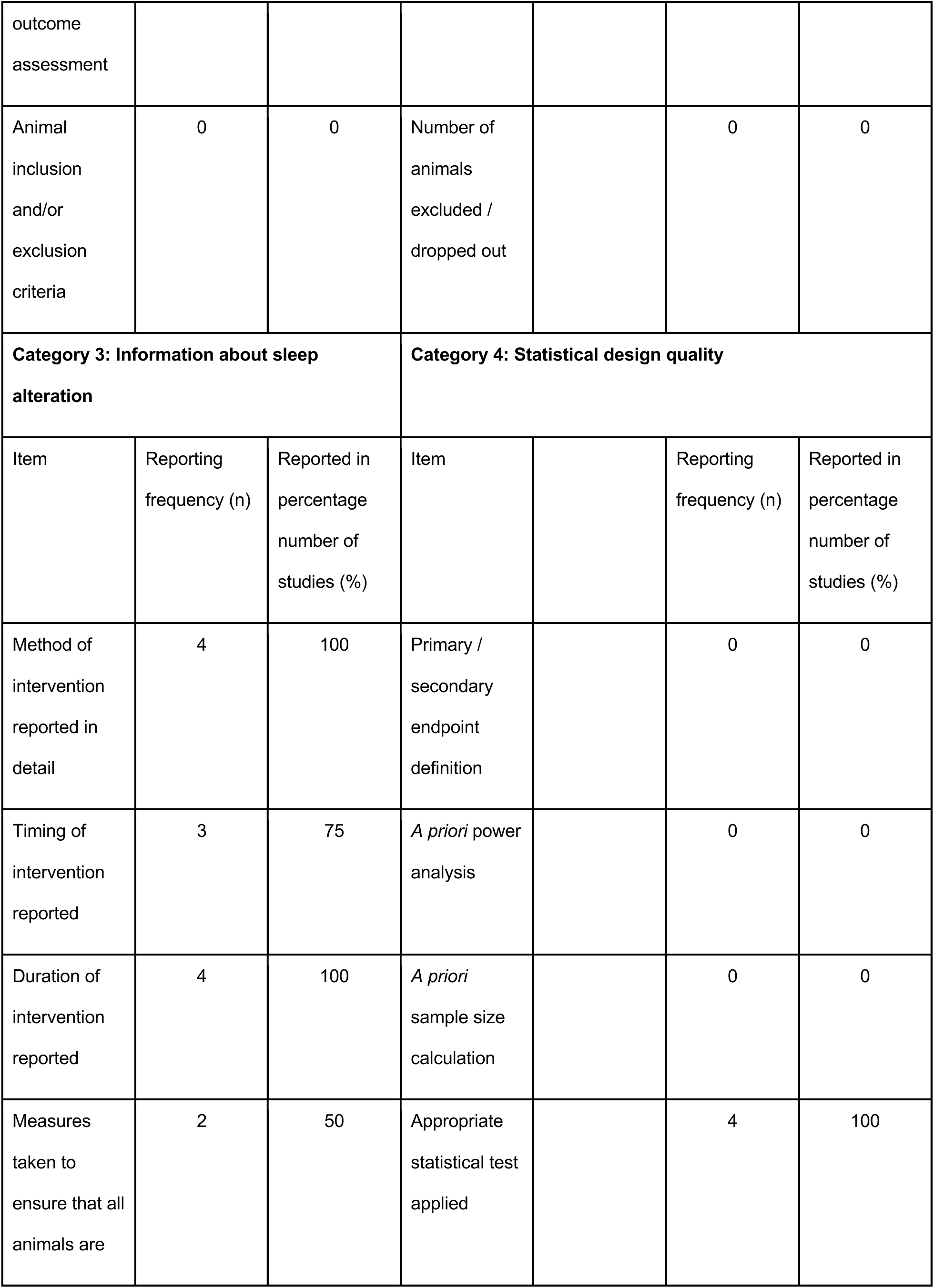
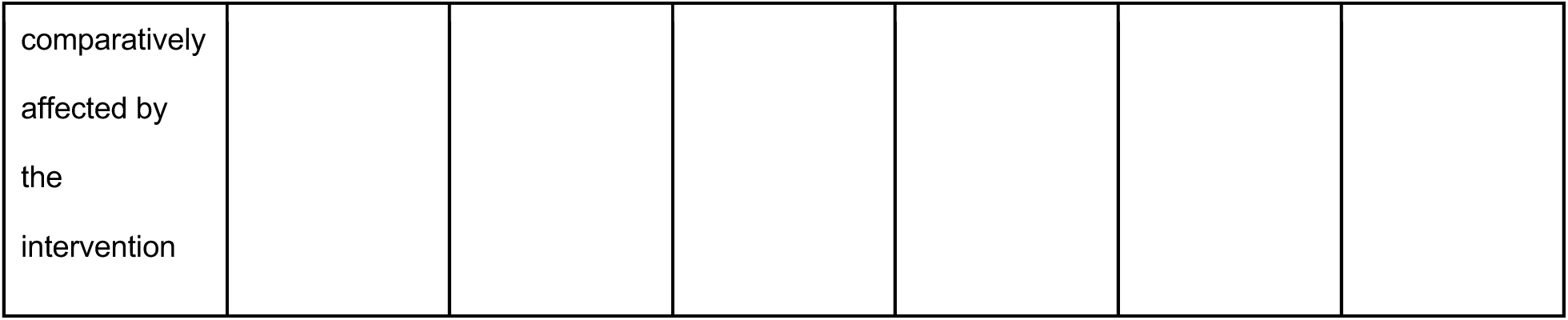
Assessment of reporting of bias-mitigating items in preclinical studies. Reporting was assessed across four categories comprising four items each. For each item, a score of 0 was assigned if it was not reported, and 1 if it was reported, yielding a total reporting score of 0-16 for each study. Values are presented as the number of preclinical studies reporting each item (n, out of four) and the corresponding percentage (%).

## 4. Discussion

OSA emerged as the only sleep disorder amongst those examined to demonstrate consistent alterations in glymphatic-related metrics across human studies. Convergent findings across distinct metric classes suggest that OSA may affect perivascular fluid dynamics through several pathways, rather than a single dominant mechanism. Meta-analysis identified a moderate inverse association between OSA severity, indexed by the AHI, and the DTI-ALPS index. This was supported by severity-stratified analyses, in which the largest pooled effect was observed in severe OSA, and by an exploratory moderate-severe OSA analysis. However, these findings should be interpreted cautiously given the small number of contributing studies. In contrast, no statistically significant association was identified between SD and glymphatic-related measures. Available evidence of NT1 similarly did not demonstrate glymphatic-related alterations, although the small number of studies limits interpretation.

### 4.1 Mechanistic interpretation

#### 4.1.1 OSA

A schematic model of physiological glymphatic transport is shown in Figure 5A. Several mechanisms may contribute to the observed disruption in glymphatic system function in OSA (Figure 5B). First, obstructive events generate large intrathoracic pressure swings, with downstream effects on intracranial pressure and cerebrovascular pulsatility^24,25^. Perivascular CSF transport is tightly coupled to cardiorespiratory oscillations and arterial pulsatility, suggesting that repeated pressure swings could disrupt perivascular exchange^6,26,27^, which may explain the observed alterations in BOLD-CSF decoupling strength in OSA^28^. Over longer timescales, OSA is associated with the development of sustained hypertension and increased arterial stiffness^29,30^. Chronic hypertension is associated with impaired glymphatic transport in spontaneously hypertensive rats^31^. OSA may therefore influence perivascular fluid dynamics through both acute pressure fluctuations and longer-term vascular dysfunction.

**Figure 5.**
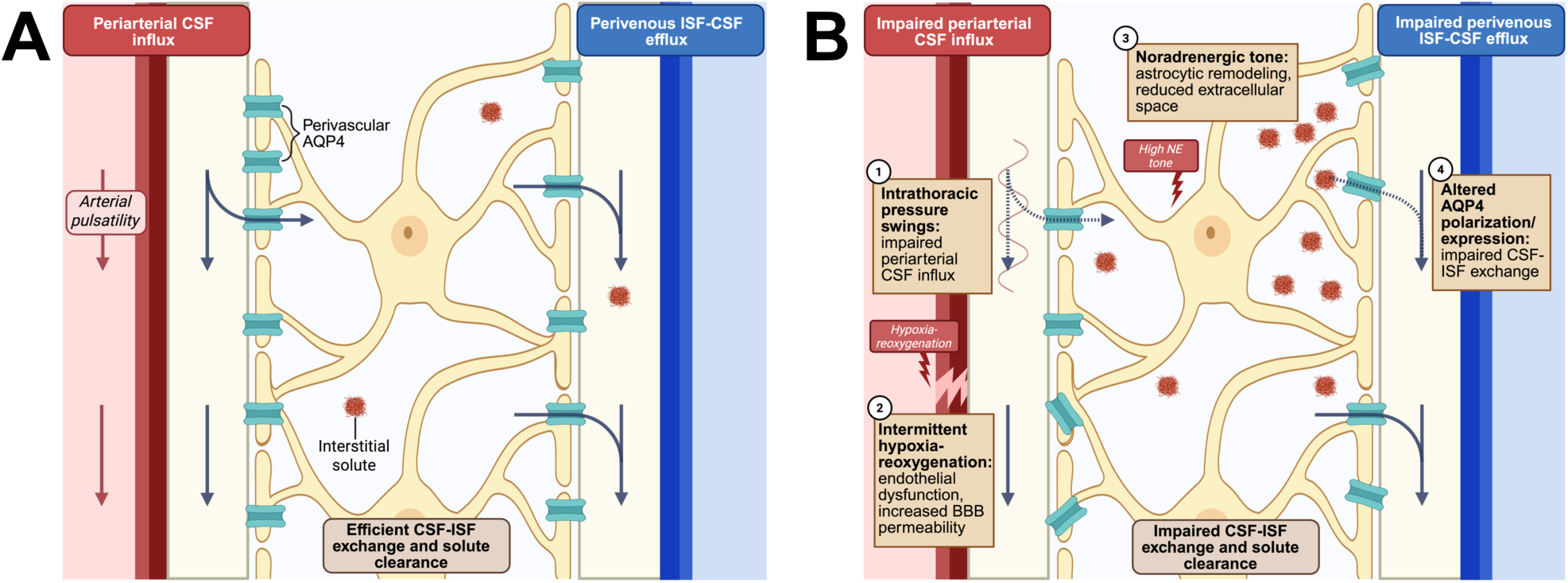
Glymphatic transport under physiological conditions and proposed mechanisms of disruption in obstructive sleep apnea (OSA). A) Under physiological conditions, arterial pulsatility supports periarterial cerebrospinal fluid (CSF) influx, enabling CSF-interstitial fluid (ISF) exchange and perivenous solute clearance. B) Proposed mechanisms of glymphatic dysfunction in OSA include (1) hemodynamic and vascular dysfunction, including acute pressure swings and chronic hypertension, which may impair CSF influx; (2) intermittent hypercapnic hypoxia, which may promote endothelial and blood-brain barrier (BBB) dysfunction; (3) elevated noradrenergic tone, which may drive astrocytic process expansion and reduced extracellular space; and (4) altered aquaporin-4 (AQP4) polarization and expression, which may impair CSF-ISF exchange. Together, these mechanisms are proposed to reduce perivascular exchange and pervenous solute clearance. Created with BioRender.com.

Second, intermittent hypercapnic hypoxia can induce endothelial stress and activation of inflammatory responses and may promote blood-brain barrier (BBB) dysfunction^32–34^. BBB and microvascular injury could plausibly influence the permeability and fluid exchange properties of the perivascular space, potentially disrupting CSF-ISF exchange. In hypoxemic adult rats, superimposed hypercapnia alters astrocytic AQP4 expression and increases BBB permeability that may alter perivascular fluid dynamics^35^.

Third, OSA is characterized by transient surges in sympathetic nerve activity during obstructive events and elevated resting sympathetic activity during wakefulness, which may collectively influence perivascular fluid dynamics^36–38^. Noradrenergic signaling can expand astrocytic processes and reduce extracellular space volume, which may impair interstitial solute movement through extracellular pathways^39^. Consistent with this, experimental data suggest that adrenergic inhibition increases CSF influx and glymphatic transport^12,40^.

Fourth, sleep fragmentation represents a core pathophysiological OSA hallmark. Preclinical sleep-fragmentation models revealed altered AQP4 expression^41^. Given that perivascular AQP4 at astrocytic endfeet facilitates CSF influx and interstitial solute clearance^1,42^, such changes provide a plausible astroglial mechanism through which fragmented sleep could impair perivascular exchange.

#### 4.1.2 OSA severity

Across severity-stratified, exploratory moderate-severe, and correlation-based analyses, greater OSA severity was consistently associated with greater glymphatic-related alteration. The inverse association between OSA severity and DTI-ALPS index is further supported by recent longitudinal evidence from a large community-based cohort study using repeated polysomnography (PSG) and DTI-ALPS assessments over four years, in which higher AHI was consistently associated with lower DTI-ALPS index^23^. This supports the presence of a dose-response pattern with severity and response increasing over time rather than a purely cross-sectional association. In the same cohort, lower DTI-ALPS indexes were also associated with poorer cognitive performance, consistent with the possibility that altered perivascular fluid dynamics may contribute to the link between OSA and cognitive decline.

Mechanistically, increasing AHI is often accompanied by sleep fragmentation and frequent arousals, greater intermittent hypoxia-reoxygenation load, larger intrathoracic/intracranial pressure swings, and increased sympathetic tone^43,44^. Greater OSA severity may therefore amplify the mechanism proposed above, providing a plausible biological explanation for the observed inverse association. This aligns with preclinical work in which chronic sleep fragmentation has been associated with larger tracer-based clearance alterations than acute models, compatible with a cumulative burden^45^.

#### 4.1.3 Sleep deprivation

There was no statistically significant association between sleep deprivation and altered glymphatic-related metrics in humans. However, most included studies involved acute disturbance (often a single night of partial or total sleep deprivation), which may be insufficient in duration to induce measurable alterations in perivascular fluid dynamics. Thus, the available evidence may reflect limited exposure rather than the definitive absence of an effect.

Acute sleep loss may differ from chronic sleep deprivation in both magnitude and impact. Intrathecal tracer-based MRI studies demonstrate altered brain tracer enrichment in individuals with chronic poor sleep quality, indicating that sustained sleep disturbance can produce measurable changes in perivascular fluid dynamics^46^. However, even a single night of sleep deprivation increases overnight CSF Aβ40 and Aβ42 concentrations relative to normal sleep in healthy adults^47^. Although CSF amyloid-β dynamics are not direct measures of glymphatic transport, these findings may suggest that short-term sleep loss can alter relevant clearance processes. Consistent with this, preclinical sleep deprivation paradigms report reduced CSF/PVS tracer influx and reduced solute clearance^48,49^. Thus, acute sleep deprivation may induce transient alterations in glymphatic system function that may be recoverable.

### 4.2 Potential implications for dementia risk

Although the predominance of cross-sectional designs in the retrieved studies limits the causal interpretation, the overall pattern in OSA is compatible with an association between sleep-disordered breathing and altered perivascular fluid dynamics. This interpretation aligns with longitudinal observations linking OSA severity to greater amyloid burden^50,51^. There is also substantial epidemiological evidence linking OSA with increased risk of cognitive decline and dementia^18,50,52^. However, whether this link is causal or coincidental is not known. It is plausible that altered perivascular fluid dynamics may mediate increased dementia risk in OSA, but establishing such link requires both thorough investigation in relevant animal models as well as longitudinal studies in human OSA patients.

### 4.3 Potential clinical and translational implications

OSA is highly prevalent worldwide, affecting an estimated ∼1 billion adults, and remains substantially underdiagnosed^53,54^. Given the scale of exposure, even small alterations in perivascular fluid dynamics in OSA could result in meaningful population-level consequences. These findings therefore support viewing OSA as a plausible and potentially modifiable contributor to brain health, and support prioritizing improved OSA identification and treatment optimization in high-risk groups.

However, a key translational uncertainty is whether effective treatment can reverse OSA glymphatic-related alterations, and whether this would result in long-term cognitive benefit. Emerging observational evidence suggests that treatment may influence perivascular markers, with optimal continuous positive airway pressure (CPAP) adherence preserving basal ganglia PVS volumes in moderate-severe OSA patients^55^. CPAP therapy has been shown to improve several outcomes in randomized controlled trials (RCTs): daytime sleepiness, quality of life and risk of cardiovascular complications^56^. However, evidence for cognitive benefit is less consistent^57^. Surgical intervention to treat OSA has been shown to improve cognitive function in some contexts, but data remains sparse^58^.

Alongside OSA, chronic sleep deprivation is widespread at the population level^59,60^. Given the scale of the exposure, even small, sustained shifts in perivascular fluid dynamics could have implications for brain health.

### 4.4 Strengths and limitations

This is, to our knowledge, the first systematic review and meta-analysis to synthesize glymphatic-related outcomes across OSA, SD and NT1, integrating both preclinical and clinical evidence across multiple glymphatic-related metrics. We also identified evidence consistent with a severity-dysfunction relationship between OSA and altered glymphatic-related metrics, supported by emerging longitudinal work and mechanistic animal studies.

However, there are several limitations. First, the number of included studies remains small, particularly for the analysis linking OSA to cognition. Substantial heterogeneity was evident across studies in sleep disturbance severity, cohort composition, analytical approaches and glymphatic/cognitive outcomes. Sleep deprivation protocols were largely acute, limiting the generalizability of results to chronic sleep deprivation. In turn, preclinical evidence came from studies using small sample sizes and may vary across model and genetic background. Moreover, sleep fragmentation models do not fully recapitulate OSA pathobiology, limiting mechanistic understanding of the contributions of intermittent hypercapnic hypoxia and intrathoracic pressure swings. One included article was retracted following the completion of the literature search and data extraction for non-scientific reasons^61^; however, its exclusion in the leave-one-out sensitive analysis did not substantially alter the pooled effect size. Finally, screening was conducted by a single reviewer. Although strict eligibility criteria were applied, this approach may carry an increased risk of missed eligible studies ^62^.

### 4.5 Heterogeneity and measurement validity

The major source of heterogeneity is that the glymphatic-related outcomes across studies do not reflect a single physiological construct but assess different components of perivascular fluid dynamics. Tracer-based methods, while providing the most direct in vivo assessment, were limited in their use across the studies (n = 3/12 meta-analyzed clinical studies), likely due to their invasiveness^63,64^. Consequently, most included studies relied on indirect, non-invasive proxies, which are sensitive to non-glymphatic factors. The DTI-ALPS index infers glymphatic function from perivascular water diffusivity^11,65^ but is also sensitive to white matter microstructure and tract geometry^66^. OSA has been associated with diffusion MRI-detectable white matter microstructural alterations^67,68^, so OSA-related white matter injury could shift the DTI-ALPS index even without changes in perivascular transport^69^. Moreover, BOLD-CSF coupling measures coordination between hemodynamic fluctuations and CSF oscillations rather than direct clearance^14^. Structural markers, such as PVS burden and CP volume, could reflect altered downstream fluid handling, rather than real-time perivascular exchange^70^. These limitations reduce the physiological specificity of the results, but do not exclude the observation that OSA-related alterations were detected across multiple glymphatic metric classes.

Additionally, OSA cognitive outcomes were assessed using non-equivalent tests spanning global cognitive screening (MoCA), executive control (Stroop), and visuomotor performance (line-tracing), limiting domain-specific comparability across cohorts^71–73^.

### 4.6 Research priorities and future directions

A major priority for future study is interventional evidence that goes beyond correlation to test reversibility of glymphatic effects. Randomized trials of CPAP therapy and other interventions, such as surgery, could test whether treatment-induced changes in OSA burden precede improvements in glymphatic proxies, and whether these glymphatic changes also correspond with improvements in cognitive performance.

A further priority is to assess glymphatic-related metrics in humans with chronic sleep deprivation, such as insomnia, as acute sleep deprivation experiments may not capture the mechanisms and magnitude of alterations in chronic sleep loss.

Current preclinical models used to study glymphatic changes in OSA prioritize sleep fragmentation, which may under-model other physiological drivers implicated in OSA, such as intermittent hypoxia and intrathoracic/intracranial pressure swings. Next-generation models should integrate intermittent hypoxia, sleep fragmentation and intrathoracic/intracranial pressure swings, given that many current models isolate individual components^74,75^.

Finally, future progress will depend on harmonizing reporting standards across sleep phenotyping in SD, NT1 and OSA, glymphatic imaging, and cognitive batteries to improve the reproducibility of studies.

## 5. Conclusions

Our systematic review and meta-analysis identified alterations in glymphatic-related measures in OSA across human studies, with evidence of a moderate inverse association between AHI and the DTI-ALPS index. In contrast, no statistically significant association was identified between SD and glymphatic-related measures, while the limited evidence available for NT1 did not demonstrate glymphatic-related alterations. Although interpretation is constrained by substantial methodological heterogeneity and reliance on indirect proxies of glymphatic function, these findings support an association between OSA and altered perivascular fluid dynamics. Further longitudinal and interventional studies are needed to establish the temporal and causal nature of this association, determine whether treatment of OSA modifies glymphatic-related metrics, and assess whether such changes are accompanied by improvements in cognitive outcomes.

## CRediT authorship contribution statement

Ruth Whitehouse: Conceptualization, Methodology, Investigation, Data curation, Formal analysis, Visualization, Writing (original draft, review & editing). Johannes Boltze: Conceptualization, Methodology, Supervision, Writing (review & editing). Robert Huckstepp: Writing (review & editing). Tracy D. Farr: Writing (review & editing). Shen Li: Writing (review & editing).

## Study registration and data availability

Originally, the research was conducted by R.W. under the University of Warwick Undergraduate Research Support Scheme (URSS) with no specific intention of publishing the results. However, the interesting outcome changed our intention later, then aiming for publication. This is the reason why the study was not pre-registered. However, we are happy to formally declare that any element of the analysis such as objectives, inclusion and exclusion criteria etc. have not been changed or amended after the literature search was started. All data, including raw data, template data collection forms; data extracted from included studies, data used for all analyses and any other materials used in the review is available for the authors upon request.

## Acknowledgement

R.W. was supported by the University of Warwick Undergraduate Research Support Scheme (URSS).

## Data availability statement

All data, including raw data, template data collection forms; data extracted from included studies, data used for all analyses and any other materials used in the review is available from the authors upon request.

## Funding statement

Only intramural funds were used to conduct this research and to create this manuscript.

## Conflict of interest disclosure

The authors declare no existing conflict of interest in relation to this work.

## Ethics approval statement

This is a systematic review. Thus, no ethical approval was required.

## Patient consent statement

This is a systematic review. Thus, no patient consent was required.

## Permission to reproduce material from other sources

Not applicable as all material was self-created.

## Clinical trial registration number

Not applicable.

## Abbreviations

AD: Alzheimer’s disease
AHI: apnea-hypopnea index
ApoE: apolipoprotein E
AQP4: aquaporin-4 channel
BBB: blood-brain barrier
BOLD: blood oxygen level-dependent
CNS: central nervous system
CP: choroid plexus
CPAP: continuous positive airway pressure
CSF: cerebrospinal fluid
DTI-ALPS: diffusion tensor imaging along the perivascular space
EEG: electroencephalogram
EPVS: enlarged perivascular spaces
HC: healthy controls
ICV: intracranial volume
ISF: interstitial fluid
IVIM: intravoxel incoherent motion
MMSE: Mini-Mental State Examination
MoCA: Montreal Cognitive Assessment
MRI: magnetic resonance imaging
NT1: narcolepsy type 1
OSA: obstructive sleep apnea
PRISMA: Preferred Reporting Items for Systematic Reviews and Meta-Analyses
PVS: perivascular spaces
RCT: randomized controlled trial
SD: sleep deprivation
SYRCLE: Systematic Review Centre for Laboratory Animal Experimentation

## Notes

### Competing Interest Statement

The authors have declared no competing interest.

### Author Declarations

This systematic review and meta-analysis used exclusively aggregate data extracted from previously published, publicly available peer-reviewed studies. The systematic review included 20 studies, of which 12 clinical studies contributed to the quantitative meta-analyses. All original data sources are identified in the manuscript's reference list and study characteristics tables, with DOI links provided where available. No individual participant-level data or restricted-access datasets were obtained or analysed.

