## Supplementary Material for "Glymphatic dysfunction across sleep disorders and its cognitive implications"

### Supplementary methods

#### *Study selection*

A systematic search of PubMed and Web of Science was performed from inception to 21 July 2025, using terms relating to sleep disturbance and glymphatic function (Supplementary Table 1). Reference lists of relevant systematic reviews were also screened to identify additional eligible studies. Records were imported into Rayyan, where duplicates were removed using automated detection followed by manual verification.

#### *Screening strategy and eligibility criteria*

Following removal of duplicates, records were screened in three stages: (i) title screening, (ii) abstract screening of remaining records, and (iii) full text assessment of potentially eligible articles. The study selection process is summarized in the PRISMA flow diagram (Figure 1, main document). Predefined inclusion and exclusion criteria for preclinical and clinical studies were applied at each stage (Supplementary Tables 2 and 3). Title and abstract screening were performed by R.W. Full texts of potentially eligible articles were then retrieved and assessed for eligibility by R.W. Records with uncertain eligibility were discussed with J.B. and resolved by consensus.

#### *Data extraction*

Study variables were extracted into structured templates by R.W. Uncertainties were resolved by consensus with J.B. For both clinical and preclinical studies, study design, publication year, study location and sample sizes were recorded. Clinical participant characteristics included age, sex, diagnostic criteria for sleep disturbance and any reported comorbidities, whereas preclinical animal characteristics included species, sex, strain, age and weight. Sleep disturbance variables extracted included classification of the condition or model, respectively (sleep deprivation (SD), narcolepsy type 1 (NT1), obstructive sleep apnea (OSA), sleep fragmentation), the diagnostic or assessment method used, severity, duration and control conditions.

Primary outcomes were glymphatic measures, comprising tracer-based clearance metrics and MRI-derived proxies, such as the diffusion tensor imaging along perivascular spaces (DTI-ALPS) index, BOLD-CSF coupling strength, enlarged perivascular space burden, choroid plexus volume and intravoxel incoherent motion (IVIM) derived perfusion fractions. For each outcome, we extracted the metric type, descriptive statistics, measurement timepoint and brain region. Since no universally validated glymphatic metric exists and outcome reporting was expected to vary, we first extracted all available glymphatic measures from each study. We developed separate hierarchical frameworks of glymphatic proxies for both preclinical (Supplementary Table 4) and clinical (Supplementary Table 5) studies before quantitative synthesis. Proxies were ranked according to physiological plausibility and measurement validity. The hierarchies were developed by R.W. and reviewed by J.B., with the final rankings agreed by consensus. When multiple glymphatic metrics were reported within a study, the highest-ranked available metric was selected for the primary meta-analysis, irrespective of the direction or statistical significance of the result. Authors' narrative or mechanistic interpretations of sleep-glymphatic relationships were also summarized narratively. Secondary outcomes included any reported cognitive measures, with the test type, outcome metric and descriptive statistics extract.

Where studies reported outcomes at multiple post-sleep deprivation timepoints within 0-48 h, a single timepoint was selected for quantitative synthesis to avoid double-counting. If authors specified a primary or main analytical timepoint, that timepoint was extracted; otherwise, the latest available timepoint within 0-48 h post-sleep deprivation was used.

When outcomes were reported as medians with ranges or interquartile ranges, values were estimated as mean ± standard deviation using validated statistical methods[1,2]. Where studies reported multiple subgroups (e.g. by OSA severity), effect sizes were pooled to generate an overall comparison with healthy controls (HC) using standard formulas for combining groups in meta-analysis[3]. When quantitative outcomes were not reported verbally, values were extracted from figures using WebPlotDigitizer[4].

***Study quality, risk of bias, and robustness assessment***

Preclinical study reporting of bias-mitigating items was assessed using a SYRCLE-based reporting framework[5]. Four categories, each comprising four items, were scored as 1 (reported) or 0 (not reported), yielding a summary reporting count (0-16) (Table 1, main document). Category 1 (general quality assurance) included allocation concealment, randomization, blinded outcome assessment, and explicit inclusion/exclusion criteria. Category 2 (animal model information) included definition of species, sex, age defined, and reporting of dropouts/exclusions. Category 3 (sleep disturbance) included detailed description of induction method, timing of intervention, duration of intervention, and measures to ensure comparable exposure across animals. Category 4 (statistical design) included definition of endpoints, a priori power analysis, a priori sample size calculation and application of an appropriate statistical test. The risk-of-bias assessment was performed by R.W. against predefined criteria. Uncertainties were resolved by consensus with J.B.

Publication bias and small-study effects were examined for meta-analyses ≥5 contributing studies by visual inspection of funnel plots and by Egger's regression test [6]. A p-value $<0.05$ on Egger's regression test was considered indicative of funnel plot asymmetry. Given the limited power of asymmetry tests with small numbers of studies, these results were interpreted with caution, particularly when less than 10 studies were available[7].

The robustness of key pooled estimates was evaluated using leave-one-out sensitivity analyses, sequentially omitting each study and recalculating the summary effect. Substantial changes in the statistical significance of the pooled effect were taken to indicate that the overall effect was strongly influenced by one or a small number of studies.

Following completion of the literature search and data extraction, the article by Zhao et al.[8] was retracted because its findings were derived from data obtained without authorization or approval from the original data owners. Because the study met the prespecified eligibility criteria and was not retracted when the search and data extraction were conducted, it was retained in the primary analysis.

***Statistical analysis***

Glymphatic and cognitive outcomes were summarized as standardized mean differences (SMDs) with 95% confidence intervals (CIs), using Hedges' *g*, to allow pooling of data reported on different scales[9]. Effect sizes compared sleep disturbance groups with HC. Outcomes were aligned such that negative values indicated impaired glymphatic or cognitive function in the sleep disturbance group relative to the HC. Where necessary, scales were reversed (i.e. multiplied by -1) to ensure a consistent direction of effect across heterogeneous glymphatic and cognitive metrics. To minimize exclusion of studies from the meta-analysis due to incomplete reporting, we attempted to obtain any missing data directly from study authors by emailing the corresponding authors of the respective eligible studies (n=3). However, no responses including additional data or clarification were received. Therefore, studies without sufficient data to compute an effect size were excluded from the meta-analysis but were retained for mechanistic synthesis.

Random-effects meta-analyses were conducted throughout, with between-study variance estimated using restricted maximum likelihood (REML) and CIs calculated using the Hartung-Knapp-Sidik-Jonkman (HKSJ) method. The HKSJ method was selected as it provides more reliable CI calculation in the presence of between-study heterogeneity and small sample sizes, both present within the analyses[10,11]. Statistical heterogeneity was quantified using the $I^2$ statistic, with 25%, 50% and 75% interpreted as low, moderate and high heterogeneity[12]. All meta-analyses, effect size calculations, heterogeneity assessments, leave-out-one sensitivity analyses, Egger's regression tests, and forest and funnel plots were conducted using Review Manager (RevMan;

version 8.1.1)[13]. The correlation between DTI-ALPS index and AHI was analyzed using JASP (version 0.95.0)[14]. A p-value <0.05 was considered statistically significant.

The primary analysis compared glymphatic outcomes between sleep disturbance groups and HC, stratified by sleep disturbance type (OSA and SD). Sub-analyses examined glymphatic outcomes across OSA severity categories and the association between DTI-ALPS index and AHI. The secondary analyses compared cognitive outcomes between sleep disturbance groups and HC, where cognitive data were available.

### Supplementary material reference list

1. Wan X, Wang W, Liu J, Tong T. Estimating the sample mean and standard deviation from the sample size, median, range and/or interquartile range. *BMC Med Res Methodol.* 2014;14:135.
2. Luo D, Wan X, Liu J, Tong T. Optimally estimating the sample mean from the sample size, median, mid-range, and/or mid-quartile range. *Stat Methods Med Res.* 2018;27(6):1785-1805.
3. Borenstein M, Hedges LV, Higgins JP, Rothstein HR. *Introduction to meta-analysis.* John wiley & sons; 2021.
4. *WebPlotDigitizer* [computer program]. Version 5.2: Automeris; 2024.
5. Hooijmans CR, Rovers MM, de Vries RB, Leenaars M, Ritskes-Hoitinga M, Langendam MW. SYRCLE's risk of bias tool for animal studies. *BMC Med Res Methodol.* 2014;14:43.
6. Egger M, Davey Smith G, Schneider M, Minder C. Bias in meta-analysis detected by a simple, graphical test. *Bmj.* 1997;315(7109):629-634.
7. Sterne JA, Egger M. Regression methods to detect publication and other bias in meta-analysis. *Publication bias in meta‐analysis: Prevention, assessment and adjustments.* 2005:99-110.
8. Zhao W, Rao J, Wang R, et al. Test-retest reliability of coupling between cerebrospinal fluid flow and global brain activity after normal sleep and sleep deprivation. *Neuroimage.* 2025;309:121097.
9. Hedges LV. Distribution Theory for Glass's Estimator of Effect size and Related Estimators. *Journal of Educational Statistics.* 1981;6(2):107-128.
10. IntHout J, Ioannidis JP, Borm GF. The Hartung-Knapp-Sidik-Jonkman method for random effects meta-analysis is straightforward and considerably outperforms the standard DerSimonian-Laird method. *BMC Med Res Methodol.* 2014;14:25.

11. Langan D, Higgins JPT, Jackson D, et al. A comparison of heterogeneity variance estimators in simulated random-effects meta-analyses. *Res Synth Methods.* 2019;10(1):83-98.
12. Higgins JP, Thompson SG, Deeks JJ, Altman DG. Measuring inconsistency in meta-analyses. *Bmj.* 2003;327(7414):557-560.
13. *Review Manager (RevMan)* [computer program]. Version 8.1.1: The Cochrane Collaboration; 2024.
14. *JASP* [computer program]. Version 0.95.0: JASP Team; 2025.
15. Lin S, Lin X, Chen S, et al. Association of MRI Indexes of the Perivascular Space Network and Cognitive Impairment in Patients with Obstructive Sleep Apnea. *Radiology.* 2024;311(3):e232274.
16. Lee HJ, Lee DA, Shin KJ, Park KM. Glymphatic system dysfunction in obstructive sleep apnea evidenced by DTI-ALPS. *Sleep Med.* 2022;89:176-181.
17. Xiong Z, Bai M, Wang Z, et al. Resting-state fMRI network efficiency as a mediator in the relationship between the glymphatic system and cognitive function in obstructive sleep apnea hypopnea syndrome: Insights from a DTI-ALPS investigation. *Sleep Med.* 2024;119:250-257.
18. Hu P, Yuan Y, Zou Y, et al. Alterations in the DTI-ALPS index and choroid plexus volume are associated with clinical symptoms in participants with narcolepsy type 1. *Sleep Med.* 2024;124:471-478.
19. Custer RM, Lynch KM, Barisano G, et al. Effects of one-night partial sleep deprivation on perivascular space volume fraction: Findings from the Stockholm Sleepy Brain Study. *Sleep Med.* 2025;131:106537.
20. Kim J, Lee HJ, Lee DA, Park KM. Choroid plexus enlargement in patients with obstructive sleep apnea. *Sleep Med.* 2024;121:179-183.
21. Ke S, Luo T, Ding Y, et al. Does Obstructive sleep apnea mediate the risk of cognitive impairment by expanding the perivascular space? *Sleep Breath.* 2025;29(2):130.
22. Eide PK, Ringstad G. Cerebrospinal fluid egress to human parasagittal dura and the impact of sleep deprivation. *Brain Res.* 2021;1772:147669.
23. van Heese EM, Gool JK, Lammers GJ, et al. MRI-based surrogates of brain clearance in narcolepsy type 1. *J Sleep Res.* 2025;34(6):e14479.
24. Eide PK, Vinje V, Pripp AH, Mardal KA, Ringstad G. Sleep deprivation impairs molecular clearance from the human brain. *Brain.* 2021;144(3):863-874.
25. Li Y, Lin S, Guo Z, et al. Decoupling of global signal and cerebrospinal fluid inflow is associated with cognitive decline in patients with obstructive sleep apnoea. *Sleep Med.* 2025;129:330-338.
26. Rane Levendovszky S, Flores J, Peskind ER, Václavů L, van Osch MJ, Iliff J. Preliminary investigations into human neurofluid transport using multiple novel non-contrast MRI methods. *J Cereb Blood Flow Metab.* 2024;44(12):1580-1592.
27. Vinje V, Zapf B, Ringstad G, Eide PK, Rognes ME, Mardal KA. Human brain solute transport quantified by glymphatic MRI-informed biophysics during sleep and sleep deprivation. *Fluids Barriers CNS.* 2023;20(1):62.

**Supplementary Table 1.** Search strategy used in PubMed and Web of Science.

| **Databases** | **Boolean Operator** |
|---|---|
| PubMed; Web of Science | (“Sleep Apnea, Obstructive” [MeSH] OR “OSA” OR “Narcolepsy” [MeSH] OR “hypersomnia” OR “central hypersomnia” OR “excessive daytime sleepiness” OR “sleep deprivation” [MeSH] OR “reduced sleep duration” OR “insufficient sleep duration”)<br>AND<br>(“Glymphatic system” [MeSH] OR “glymphatic pathway” OR “cerebrospinal fluid” OR “CSF dynamics” OR “CSF circulation” OR “CSF drainage” OR “perivascular clearance” OR “lymphatic drainage of the brain”)<br>Date of search: 21 July 2025<br>No filters applied. |

**Supplementary Table 2.** Preclinical inclusion and exclusion criteria for systematic review. For studies meeting more than one exclusion criterion, the study was assigned to a single criterion in the PRISMA flow diagram.

| **Inclusion** | **Exclusion** |
|---|---|
| Animal studies that experimentally induce or assess sleep disturbances, including sleep deprivation, Obstructive sleep apnea (OSA) models, or narcolepsy models. | Animal studies that do not assess both sleep disturbance and glymphatic or CSF-related outcomes. |
| Studies that quantitatively evaluate glymphatic system function, including CSF clearance, perivascular clearance or glymphatic drainage. | Studies assessing only sleep-related outcomes or only CSF clearance/glymphatic outcomes in isolation. |
| Original research articles. | Review articles, meta-analyses, conference abstracts, editorials, case reports, or protocols. |
| Publications available in English. | Publications in languages other than English or not available in English. |
| Full text access available. | Full text access unavailable. |

**Supplementary Table 3.** Clinical inclusion and exclusion criteria for systematic review. For studies meeting more than one exclusion criterion, the study was assigned to a single criterion in the PRISMA flow diagram.

| **Inclusion criteria** | **Exclusion criteria** |
|---|---|
| Studies involving human participants with documented sleep disturbances, including Obstructive sleep apnea (OSA), narcolepsy or hypersomnia, reduced sleep duration. | Studies that do not assess both sleep disturbance and glymphatic or CSF clearance-related outcomes. |
| Studies that evaluate glymphatic system function, CSF clearance, or perivascular clearance using quantitative imaging techniques or biomarker/tracer investigations. | Studies that do not report glymphatic-related outcomes. |
| Original research articles. | Review articles, meta-analyses, conference abstracts, editorials, case reports or protocols. |
| Publications available in English | Publications in languages other than English or not available in English. |
| Full text access available. | Full text access unavailable. |

**Supplementary Table 4.** Hierarchy of preclinical glymphatic outcome measures used for quantitative synthesis. Higher ranks indicate higher priority for inclusion when multiple glymphatic-related metrics were reported in a single study. The highest-ranked available metric was selected as the primary outcome for meta-analysis.

| **Rank** | **Glymphatic construct** | **Primary outcome metric** | **Secondary/ fallback metric(s)** |
|---|---|---|---|
| 1 | *Parenchymal clearance kinetics* | Frontal cortex interstitial fluid (ISF) clearance: % injected dose (%ID) cleared from cortex at 90 min ($^{125}$I-apoE3) | Frontal cortex ISF clearance: %ID cleared at 90 min ($^{14}$C-inulin) |
| 2 | *Downstream lymphatic outflow (deep cervical lymph nodes)* | Normalized tracer fluorescence in deep cervical lymph nodes (dCLNs), expressed as percentage of injected dose (%) | Normalized area fraction of tracer-positive signal in dCLNs; normalized fluorescence / area ratio |
| 3 | *Parenchymal tracer burden (single timepoint)* | Mean parenchymal fluorescence intensity at 60 min in frontal cortex (e.g. 100 µm sections) | |
| 4 | *Perivascular pathway signal* | Perivascular space (PVS) fluorescence intensity at 60 min (e.g. along penetrating arteries; 100 µm sections) | |
| 5 | *Regional parenchymal intensity* | Normalized fluorescence cortex (%) | Normalized fluorescence hippocampus (%) |

**Supplementary Table 5.** Hierarchy of clinical glymphatic outcome measures used for quantitative synthesis. Higher ranks indicate higher priority for inclusion when multiple glymphatic-related metrics were reported in a single study. The highest-ranked available metric was selected as the primary outcome for meta-analysis.

| Rank | Glymphatic construct | Primary outcome metric | Secondary/ fallback metric(s) |
|---|---|---|---|
| 1 | *Tracer-based late-phase clearance* | Late-phase tracer retention, brain-wide (e.g. 24-48 h post-injection; MRI or PET) | If whole-brain unavailable: (i) white-matter or subcortical ROIs, then (ii) cortical ROIs |
| 2 | *Meningeal lymphatic outflow* | Tracer enrichment in parasagittal dura at late timepoint (e.g. 24 h) | – |
| 3 | *Upstream CSF reservoir* | Tracer enrichment in subarachnoid CSF at late timepoint (e.g. 24 h) | – |
| 4 | *Diffusion-based structural glymphatic proxy* | DTI-ALPS index, bilateral mean | Unilateral or regional ALPS indices |
| 5 | *Functional CSF-brain coupling (dynamic)* | BOLD-CSF coupling strength using global or downstream bold signal (gBOLD or dBOLD) | aBOLD-based CSF-BOLD coupling metrics |
| 6 | *Perivascular space burden* | PVS volume fraction in centrum semiovale (intracranial volume-corrected preferred) | EPVS regional scores (e.g. hippocampus, midbrain) |
| 7 | *Interstitial fluid content proxy* | Free Water Index (diffusion MRI) | – |
| 8 | *Choroid plexus morphology* | Choroid plexus volume intracranial-volume-corrected | Raw choroid plexus volume |
| 9 | *Model-derived transport metrics* | Advection/flow speed (brain-wide mean; if unavailable, regional mean at 24-48 h) | Fluid influx rate (24-48 h) |
| 10 | *Vascular / perfusion and exchange proxies* | Pseudodiffusion coefficient D* (IVIM) | – |

**Supplementary Table 6.** Supplementary study characteristics of included studies. Abbreviations: HC, healthy controls; SD, sleep disturbance; PSG, polysomnography; MRI, magnetic resonance imaging; DTI-ALPS, diffusion tensor image analysis along the perivascular space; OSA, obstructive sleep apnea; AHI, apnea-hypopnea index; fMRI, functional magnetic resonance imaging; NR, not reported; ICSD-3, International Classification of Sleep Disorders, Third Edition; MSLT, Multiple Sleep Latency Test; CSF, cerebrospinal fluid; hcrt-1, hypocretin-1; PSD, partial sleep deprivation; TSD, total sleep deprivation; SF, sleep fragmentation; EEG, electroencephalogram; EMG, electromyography; FITC, fluorescein isothiocyanate; AQP4, aquaporin-4; $^{125}I$, iodine-125; apoE, apolipoprotein E; $^{14}C$, carbon-14.

| Author, year | Species/strain | Country | Sleep disturbance defined/identified | Follow-up time points | Glymphatic imaging/ tracer details | Duration/ severity of sleep disturbance |
|---|---|---|---|---|---|---|
| Lin et al., 2024[15] | — | China | PSG | Baseline | MRI; DTI-ALPS | Mild to severe OSA (AHI) |
| Lee et al., 2022[16] | — | Korea | PSG | Baseline | MRI; DTI-ALPS | Mild to severe OSA (AHI) |
| Xiong et al., 2024[17] | — | China | PSG | Baseline | MRI; DTI-ALPS index | Moderate to severe OSA (AHI) |
| Zhao et al., 2025[8] | — | China | Actigraphy/diaries/questionnaires plus PSG/oximetry pre-study screening; in-lab monitoring during protocol | After TSD | Resting-state fMRI | Acute TSD, 24 h |
| Hu et al., 2024[18] | — | China | Diagnosis based on ICSD-3; MSLT; CSF hcrt-1 assays | Baseline | MRI; DTI-ALPS | Moderate to severe; disease duration: 5.54 ± 3.09 years. |
| Custer et al., 2025[19] | — | Sweden | At-home PSG | Evening after sleep condition | Structural MRI | Acute PSD, 24 h (~3 h sleep) |
| Kim et al., 2024[20] | — | Korea | Lab PSG | Baseline | Structural MRI | Mild to severe OSA (AHI) |
| Ke et al., 2025[21] | — | China | Lab PSG | Baseline | Structural MRI | Mild to severe (AHI) |
| Eide and Ringstad, | — | Norway | Protocol-driven TSD | 6-48 h post-injection | Gadobutrol intrathecal CSF tracer; structural MRI | Acute TSD, 24 h |

| | | | | | | |
|---|---|---|---|---|---|---|
| 2021[22] | | | | | | |
| van Heese et al., 2025[23] | — | The Netherlands | Diagnosis based on ICSD-3; lab PSG; MSLT; CSF hcrt-1 assays | Baseline | MRI; DTI-ALPS | Moderate to severe; disease duration: median 83 months |
| Eide et al., 2021[24] | — | Norway | Wakefulness verified by staff; sleep group recorded hours slept and subjective sleep quality | 1.5-48 h | Gadobutrol intrathecal CSF tracer; structural MRI | Acute TSD, 24 h |
| Li et al., 2025[25] | — | China | Overnight PSG | Baseline | Resting-state fMRI | Mild to severe OSA (AHI) |
| Rane Levendovsky et al., 2024[26] | — | USA | Hourly phone check-ins | Morning (08:00-10:00) following sleep condition | Diffusion MRI | Acute TSD, 24 h |
| Vinje et al., 2023[27] | — | Norway | Protocol-driven TSD | 0-48 h post-injection | Structural MRI; Intrathecal gadobutrol CSF tracer | Acute TSD, 48 h |
| Roy et al., 2022[28] | — | USA | Overnight PSG | Baseline | MRI; DTI-ALPS | Moderate to severe OSA (AHI) |
| Gumeler et al., 2023[29] | — | Turkey | Diagnosis based on ICSD-3; night PSG; MSLT | Baseline | MRI; DTI-ALPS | NR |
| Deng et al., 2024[30] | Mouse; C57BL/6 | China | Protocol-driven SF via orbital shaker (EEG/EMG-verified) | Within 24 h of final SF intervention | Cisterna magna 40 kDa FITC-dextran; fluorescence microscopy of brain sections (tracer influx). | Acute SF, 3 days; chronic SF, 30 days; severe SF (~30 arousals/h) during light phase |
| Vasciaveo et al., 2023[31] | Mouse; 5xFAD and B6SJL | Italy | Protocol-driven SF via tilting platform (EEG/EMG-verified) | Post-intervention | No CSF tracer/glymphatic imaging; AQP4 used as proxy marker | Chronic SF (30 days); severe SF (~20 arousals/h) 24 h/day |
| Achariyar et al., 2016[32] | Mouse; C57BL/6J | USA | Protocol-driven sleep deprivation (6-9 AM); wakefulness maintenance protocol | Post-intervention | ISF clearance assessed by intracortical $^{125}$I-apoE and $^{14}$C-inulin | Acute PSD, 3 h |
| Liu et al., 2017[33] | Mouse; C57BL/6J | China | Protocol-driven sleep deprivation through modified multiple platform method (48 h) | Immediate post-intervention (10-60 min) | Cisterna magna injection of FITC-dextran (70 kDa) with in vivo two-photon microscopy to quantify perivascular | Acute TSD, 48 h |

| | | | | | |
|---|---|---|---|---|---|
| | | | | | (PVS) CSF influx |
